# Socioeconomic inequalities in COVID-19 infection and vaccine uptake among children and adolescents in Catalonia, Spain

**DOI:** 10.1101/2023.10.17.23297134

**Authors:** Irene López-Sánchez, Aida Perramon, Antoni Soriano-Arandes, Clara Prats, Talita Duarte-Salles, Berta Raventós, Elena Roel

## Abstract

**IMPORTANCE:** The relationship between socioeconomic deprivation and COVID-19 infection and vaccination among children and adolescents remains unclear.

**OBJECTIVE:** To investigate the association between deprivation and COVID-19 vaccine uptake and infection among children and adolescents before and after the vaccination rollout in Catalonia, Spain.

**DESIGN AND SETTING:** Population-based cohort study using primary care electronic health records from the Information System for Research in Primary Care. Individuals were followed 3 months before the start of the vaccination campaign in Spain and 3 months after the vaccination rollout among adolescents and children.

**PARTICIPANTS:** Children (5-11 years) and adolescents (12-15 years) with at least 1 year of prior history observation available and without missing deprivation data.

**EXPOSURE:** Deprivation, assessed using an ecological socioeconomic deprivation index (SDI) score for census tract urban areas and categorized into quintiles.

**MAIN OUTCOMES AND MEASURES:** COVID-19 infection and COVID-19 vaccination. For each outcome, we calculated cumulative incidence and crude Cox proportional-hazard models by SDI quintiles, and estimated hazard ratios (HRs) of COVID-19 infection and vaccine uptake relative to the least deprived quintile, Q1.

**RESULTS:** Before COVID-19 vaccination rollout, 290,625 children and 179,685 adolescents were analyzed. Increased HR of deprivation was associated with a higher risk of COVID-19 infection in both children (Q5: 1.55 [95% CI, 1.47 - 1.63]) and adolescents (Q5: 1.36 [95% CI, 1.29 - 1.43]). After the rollout, this pattern changed among children, with lower risk of infection in more deprived areas (Q5: 0.62 [95% CI, 0.61 - 0.64]). Vaccine uptake was higher among adolescents (72.6%) than children (44.8%), but in both age groups, non-vaccination was more common among those living in more deprived areas (39.3% and 74.6% in Q1 vs. 26.5% and 66.9% in Q5 among children and adolescents, respectively).

**CONCLUSIONS AND RELEVANCE:** In this cohort study, children and adolescents living in deprived areas were at higher risk of COVID-19 non-vaccination. Socioeconomic disparities in COVID-19 infection were also evident before vaccine rollout, with a higher infection risk in deprived areas across age groups. Our findings suggest that changes in the association between deprivation and infections among children after the vaccine rollout were likely due to testing disparities.

## Introduction

The coronavirus disease 2019 (COVID-19) pandemic disproportionately affected vulnerable populations with low socioeconomic status (SES)^1^. Although extensive research has already been conducted on this topic^2–7^, the relationship between SES and COVID-19 infection and vaccination among children and adolescents remains unclear.

Research from Canada and the United Kingdom (UK) suggests that children and adolescents with low SES face a higher risk of low COVID-19 vaccine uptake compared to their high SES peers^8, 9^. Studies from the United States (US) also show that children from low-income households are more susceptible to SARS-CoV-2 infection^10, 11^. However, generalizing these findings to other regions may not be applicable, and evidence on whether vaccination campaigns have impacted this association is scarce.

In Spain, evidence of socioeconomic inequalities in COVID-19 vaccine uptake and infection is limited to adults. A study by Roel et al. found a pattern of increased HR infection with deprivation among adults aged 40 years and above which decreased after vaccination rollout^7^. Identifying patterns of socioeconomic disparities in COVID-19 vaccination and infection in Catalonia (Spain) could provide key information to guide immunization strategies in Spain and other countries where vaccines are readily accessible. In this study, we aimed to investigate the association between deprivation (a proxy measure of SES) and COVID-19 infection 3 months before the COVID-19 vaccine rollout and 3 months after the start of vaccination of children and adolescents living in urban areas of Catalonia, Spain. We also investigated the associations between deprivation and COVID-19 vaccine uptake among children and adolescents.

## Methods

### Study Design and Data Source

We conducted a population-based cohort study using primary care data from the Information System for Research in Primary Care (SIDIAP; www.sidiap.org) database, which contains pseudo-anonymized electronic health records of approximately 75% of the Catalan population and is representative in geography, age, and sex^12^.

SIDIAP includes information on sociodemographics, disease diagnoses, laboratory tests, medication use and death^12^. In addition, this database has been linked to the Catalan public health vaccine registry and to a population-based register of hospital discharge records from public and private hospitals of Catalonia (Conjunt Mínim Bàsic de Dades d’Alta Hospitalària, CMBD-AH)^13^. It has been standardized to the Observational Medical Outcomes Partnership (OMOP) Common Data Model (CDM), and the provenance of the data, specifically for capturing COVID-19-related outcomes, has been well documented^14^.

This project was approved by the Clinical Research Ethics Committee of the IDIAPJGol (project code: 21/052-PCV).

### Study periods

We followed children aged 5 to 11 years and adolescents aged 12 to 15 years during two time periods 1) a pre-vaccination period, three months before the start of the vaccination rollout in Spain, from the 26th of September to the 26th of December 2020^15^; and 2) a vaccination period, three months after the start of the vaccination rollout among adolescents (from the 1st August to the 1st November 2021^16^) and children (from the 15th of December 2021 to the 15th of March 2022^17^). Thus, four cohorts were followed, one for each age group and time period (Figure 1), with the follow-up of all cohorts starting on the first day of each time period (index date). The vaccination period for children was set to three months, and, to ensure consistency across cohorts, the same time window was applied to the other cohorts^18, 19^.

**Figure 1.**
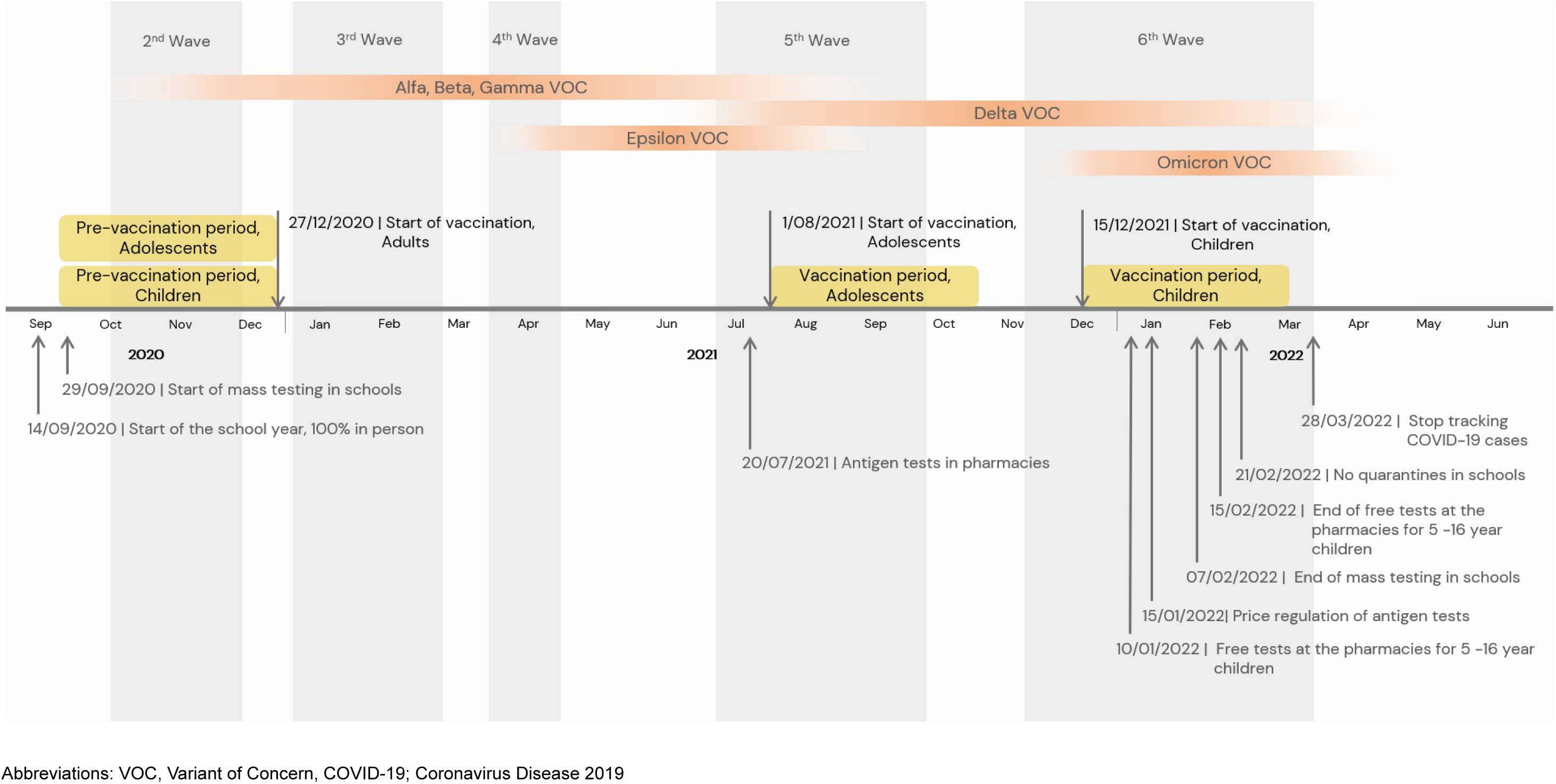
Timeline of the cohort studies.

### Setting

Our study was conducted against the backdrop of the evolving COVID-19 pandemic in Catalonia, Spain. The region experienced distinct phases of the pandemic, each marked by unique challenges and responses.

During the pre-vaccination period, Catalonia was experiencing the second wave of the COVID-19 pandemic, with incidence rates ranging from 120 to over 400 cases per 100,000 inhabitants^20, 21^. In September 2020, educational settings reopened for in-person activities after closing in March 2020 and enforced stringent non-pharmaceutical interventions to mitigate the virus transmission. These measures consisted of hand-washing, mandatory mask use for children over 5 years old, social distancing, organization of children and teachers into bubble groups to maintain the same groups of individuals and therefore facilitate contact tracing, and the improvement of ventilation of classrooms. Other public health measures such as mass testing were implemented in schools located in areas with high incidence of COVID-19 to reduce transmission^22, 23^. In the event of a confirmed case, testing of all classmates was performed and, regardless of the PCR test result, the whole group was quarantined for 10 days^24^.

In late July 2021, pharmacies began selling antigen tests following a surge of COVID-19 cases that coincided with the emergence of the Delta variant^25, 26^. When the academic year 2021-2022 started in September after a two-month break, school protocols were revised. In the event of a confirmed infection, close contacts were required to undergo testing, which was provided free of charge through the public health system. Those who had previously been infected within the last 6 months or were fully vaccinated, were directed to obtain a supervised antigen test at authorized pharmacies. Individuals who did not meet these criteria were instructed to undergo a PCR test at a primary healthcare center^27^. Between October and November 2021, COVID-19 incidence rates decreased from 1,630 to 716 cases per 100,000 inhabitants^20^.

In the vaccination period for children, the incidence of COVID-19 in Catalonia increased sharply due to the higher transmissibility rate of the Omicron variant, rising from 551 cases in December 2021 to 3,285 cases per 100,000 inhabitants in January 2022, but then rapidly decreased to 300 cases per 100,000 inhabitants by mid-March when most of the population had been infected^20, 26^. In January 2022, the government regulated the price of antigen tests to improve accessibility and offered free antigen tests to fully vaccinated children and adolescents when a positive case appeared in their social bubble^28, 29^. Testing was restricted to unvaccinated children and adolescents without prior history of infection over the last three months. This measure ended in February 2022, along with mass testing and quarantines in schools^30–32^.

### Study participants

We included all children and adolescents registered in the SIDIAP database as of 26 September 2020 (index date for all participants). We excluded those with less than 1 year of prior medical history available at the index date and those with missing data on deprivation (Figure 2). When analyzing COVID-19 infection, we excluded those individuals with an infection ≤ 6 weeks prior to the start of the study period. When analyzing COVID-19 vaccine uptake, we excluded those who had already received a COVID-19 vaccine prior to the start of the vaccination period, and those who tested positive for COVID-19 within the previous 8 weeks, since Spanish policies required 8-week delay between infection and vaccination^33^. Figure 2 shows the flow chart of inclusion and exclusion criteria.

**Figure 2.**
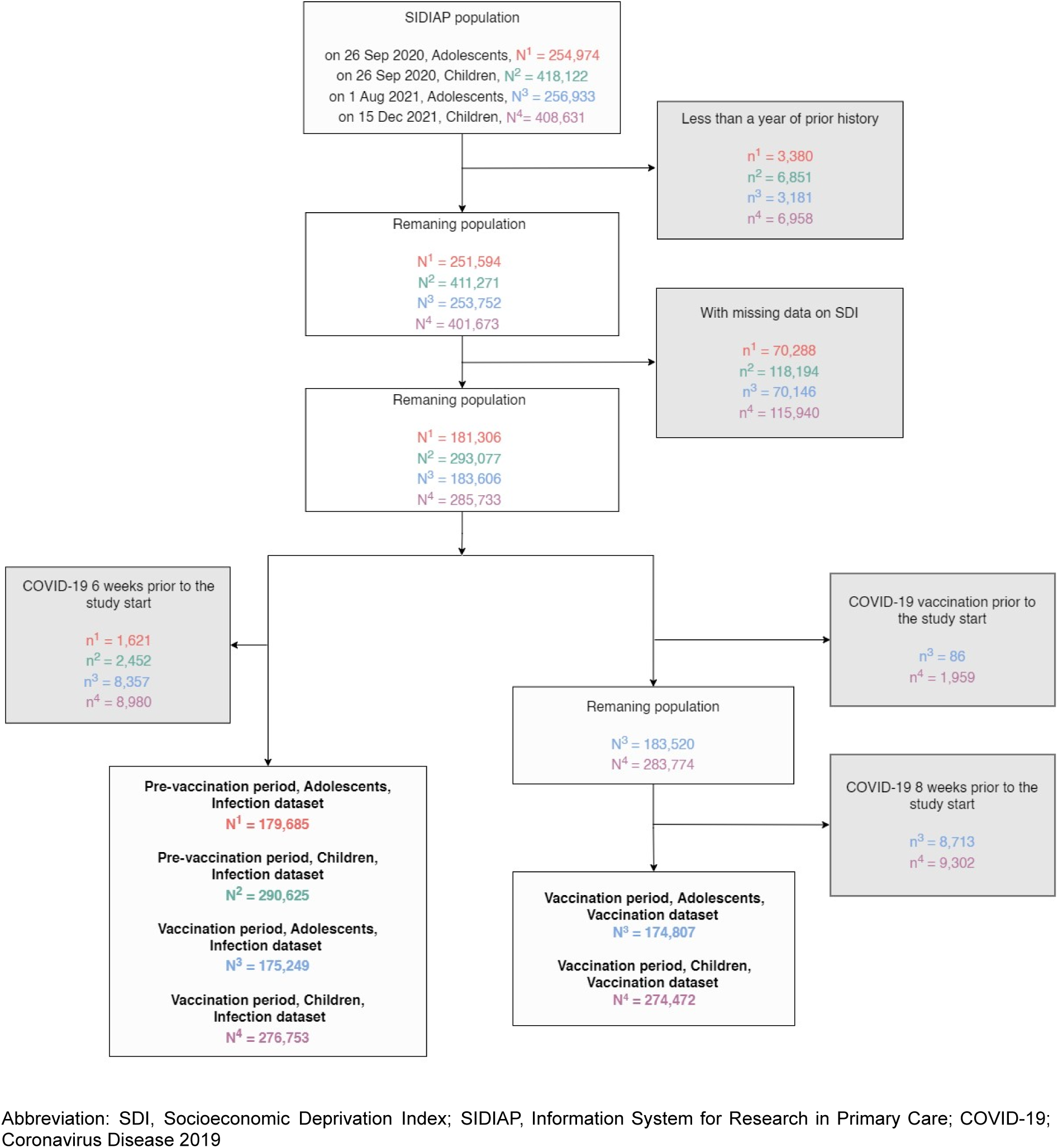
Flowchart showing the inclusion and exclusion criteria for study population in analysis of socioeconomic inequalities in COVID-19 vaccination and infection in children and adolescents, Catalonia, Spain.

For each period, children and adolescents were followed until the occurrence of the outcome of interest, end of the study period, exit from the SIDIAP database, or death, whichever occurred first. When analyzing COVID-19 vaccine uptake, participants who had been infected prior to vaccination were censored at the date of infection, since infections were competing risks for vaccination^34–36^.

### Variables

Deprivation was assessed using a Socioeconomic Deprivation Index (SDI) score based on the place of residence (a proxy of SES), the *Mortalidad en áreas pequeñas españolas y desigualdades socioeconómicas y ambientales* (MEDEA) deprivation index^37^. This index was calculated at census tract areas using data from the 2001 national census in Spain, and was only available for urban areas (municipalities with >10,000 inhabitants and a population density >150 habitants/km2). This index has been linked in SIDIAP to each individual’s most recent residential address and categorized into five quintiles, the first being the least deprived (Q1) and the fifth being the most deprived (Q5).

Our outcomes of interest were COVID-19 infection and vaccination. COVID-19 infections were defined as a positive SARS-CoV-2 test result [antigen or polymerase chain reaction (PCR)] or a clinical COVID-19 diagnosis without a record of a COVID-19 infection 6 weeks prior, with the test or diagnosis date as the date of infection^38^. In each study period, we only considered the first COVID-19 infection per person. COVID-19 vaccination was defined as having received at least one dose of a COVID-19 vaccine authorized for use in children and adolescents (BNT162b2 or mRNA-1273). The date of vaccination was the date of the first dose administration.

Additionally, we extracted age, sex, and nationality by country’s geographic region at baseline and the number of SARS-CoV-2 tests performed during follow-up.

### Statistical Analysis

We described participants’ characteristics at baseline, with counts and percentages for categorical variables and median and interquartile ranges (IQRs) for continuous variables.

We calculated the cumulative incidence of COVID-19 infection for each study period and age group of our study, according to the SDI quintile, by dividing the number of incident COVID-19 infections by the number of individuals at risk. Vaccination uptake was calculated at the end of the vaccination period by dividing the number of individuals vaccinated by the total number of individuals eligible for vaccination.

We described the proportion of individuals who had undergone one or more tests registered in the public health system during follow-up, according to the deprivation quintile at the end of each study period. To assess the association between SDI and COVID-19 infection, we performed crude Cox proportional-hazard regression models to estimate hazard ratios (HRs) with 95% confidence intervals (CI) of COVID-19 infection by deprivation quintile relative to the least deprived quintile, Q1. To assess the association between SDI and COVID-19 vaccine uptake, we performed crude cause-specific Cox proportional-hazard regression models to estimate the HR with 95% CIs of vaccination by SDI quintile relative to Q1. All the analyses were stratified by age group and study period. We visually inspected log-log survival curves to check the proportional hazard assumptions for SDI (eFigure 1 in the Supplement). We performed crude models taking into consideration our assumptions on the relationship between our exposure and our outcomes of interest, which we represented with direct acyclic graphs (DAGs) (eFigure 2 and eFigure 3 in the Supplement). As a secondary analysis, we stratified all our analyses by sex.

**Figure 3.**
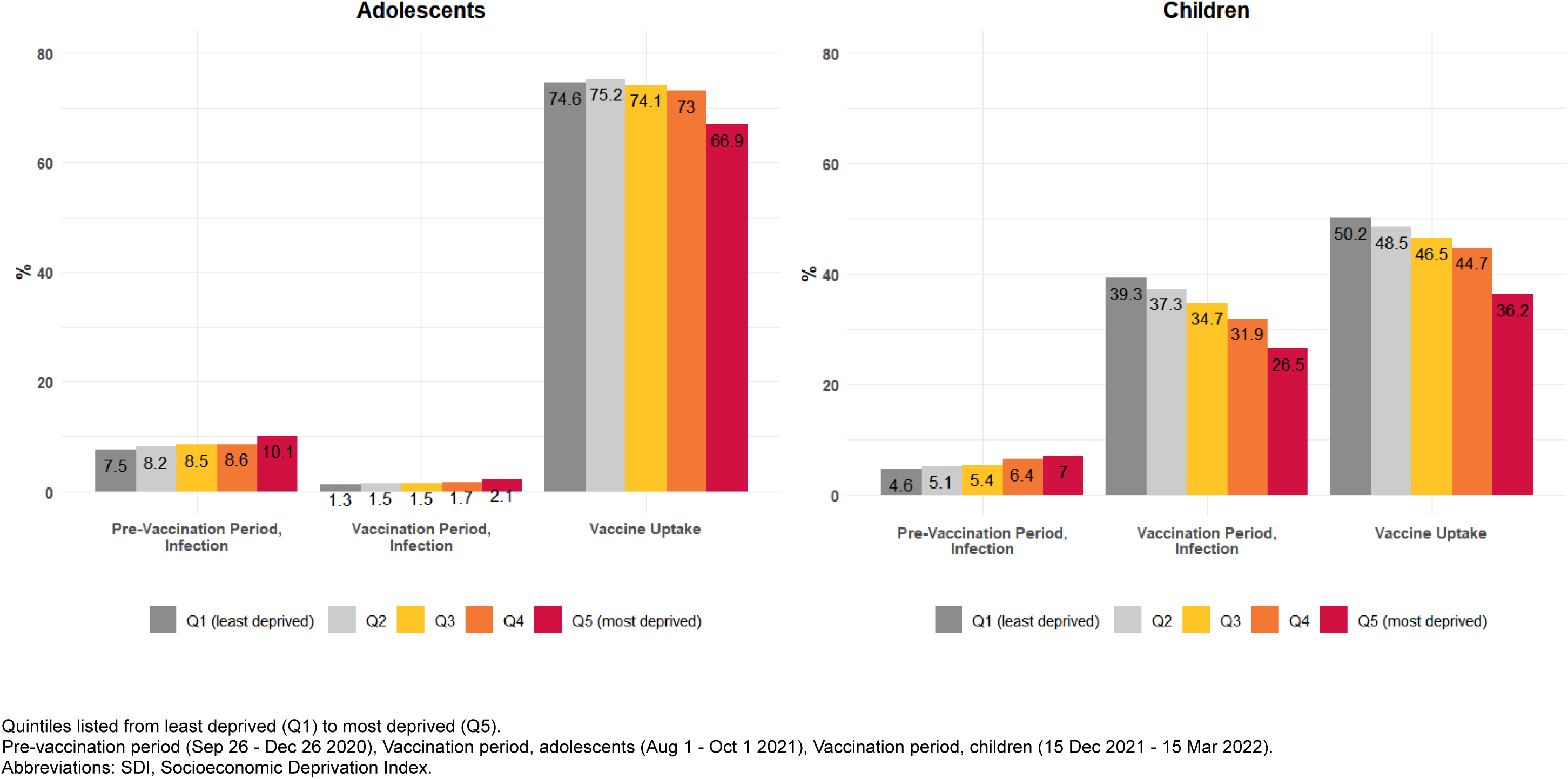
Cumulative incidence of COVID-19 infection and vaccine uptake by age group and Socioeconomic Deprivation Index quintile, before and after the start of the vaccination rollout.

As a sensitivity analysis, we excluded individuals with a prior history of COVID-19 infection at any point prior to the start of each study period, as it might have influenced the risks of infection as well as of being vaccinated.

We used R version 4.1 for data curation and analysis.

## Results

Before the vaccination rollout, 179,685 adolescents (92,442 [51.4%] male) and 290,625 children (149,605 [51.5%] male) were included in the study (Table 1). The majority of participants were Spanish (86.2% children and 88.9% adolescents), and the distribution of SDI quintiles was similar across age groups. Similar baseline characteristics were observed among children and adolescents after the vaccination rollout (eTable 1 and eTable 2 in the Supplement). Prior COVID-19 infection was low (0.6%) among children and adolescents in the pre-vaccination period, but increased to 17% in the vaccination period in both age groups (eTable 1 and eTable 2 in the Supplement).

**Table 1.** Baseline characteristics of study participants as of September 26, 2020.

| Characteristics | Children<br>(5-11 years) | Adolescents<br>(12-15 years) |
| --- | --- | --- |
| **Total No.** | 290625 | 179685 |
| **Age (median [IQR])** | 8.0 [6.0, 10.0] | 13.0 [12.0, 14.0] |
| **Sex No. (%)** |  |  |
| Female | 141020 (48.5) | 87243 (48.6) |
| Male | 149605 (51.5) | 92442 (51.4) |
| **Nationality No. (%)** |  |  |
| Spain | 250644 (86.2) | 159824 (88.9) |
| Africa | 12915 (4.4) | 5247 (2.9) |
| Asia & Oceania | 9492 (3.3) | 5065 (2.8) |
| Central & South America | 8802 (3.0) | 4908 (2.7) |
| Eastern Europe | 5487 (1.9) | 3030 (1.7) |
| Europe & North America | 3285 (1.1) | 1611 (0.9) |
| **SDI quintile<sup>a</sup> No. (%)** |  |  |
| Q1 (least deprived) | 53140 (18.3) | 34524 (19.2) |
| Q2 | 55703 (19.2) | 35637 (19.8) |
| Q3 | 56850 (19.6) | 35686 (19.9) |
| Q4 | 60099 (20.7) | 35994 (20.0) |
| Q5 (most deprived) | 64833 (22.3) | 37844 (21.1) |
| **Previous COVID-19 Infection No. (%)** | 1814 (0.6) | 1039 (0.6) |

<sup>a</sup>Quintiles listed from least deprived (Q1) to most deprived (Q5).
Abbreviations: IQR, interquartile range; SDI, Socioeconomic Deprivation Index.

### COVID-19 tests, cumulative incidence of infection & vaccine uptake

Before the vaccination rollout, more tests were performed to children and adolescents living in more deprived areas: 32.1% (Q1), vs. 37.6% (Q5) in children and 45.9% (Q1), vs 48.6% (Q5) in adolescents (eFigure 3 in the Supplement). Cumulative incidence of COVID-19 infection was 5.8% in children and 8.6% in adolescents. Infections were more frequent in more deprived areas in both age groups, showing a clear gradient of increased infection with deprivation: 4.6% (Q1), 5.4% (Q3) and 7% (Q5) in children, and 7.5% (Q1), 8.5% (Q3) and 10.1% (Q5) in adolescents (Figure 3; eTable 3 in the Supplement). After the vaccination rollout, testing was higher among those living in less deprived areas: 71.4% (Q1), vs 59.5% (Q5) in children and 17.9% (Q1), vs 14.2% (Q5) in adolescents (eFigure 3 in the Supplement). Cumulative incidence of infection was 33.6% for children and 1.6% for adolescents. The pattern of increased COVID-19 infection with increased deprivation was still seen among adolescents (1.3% [Q1], 1.5% [Q3] and 2.1% [Q5]). However, we found the opposite pattern among children, with those in the least deprived quintile having higher incidence proportion of infection (39.3% Q1, 34.7% Q3 and 26.5% Q5).

Cumulative incidence of vaccination was 44.8% for children and 72.6% for adolescents. We also found a pattern of increased vaccination among those living in less deprived areas, with an incidence proportion of 51.9% (Q1) vs. 38.3% (Q5) in children and 84.8% (Q1) vs. 75.7% (Q5) in adolescents.

### Association between SDI and COVID-19 infection and vaccine uptake

Before the vaccination rollout, we observed a pattern of increased risk of COVID-19 infection with increased deprivation in both age groups (Figure 4; eTable 4 in the Supplement). For instance, among children, HR of infection relative to Q1 ranged between 1.12 (95% CI 1.06 - 1.18) in Q2 areas and 1.55 (1.47 - 1.63) in Q5 areas.

**Figure 4.**
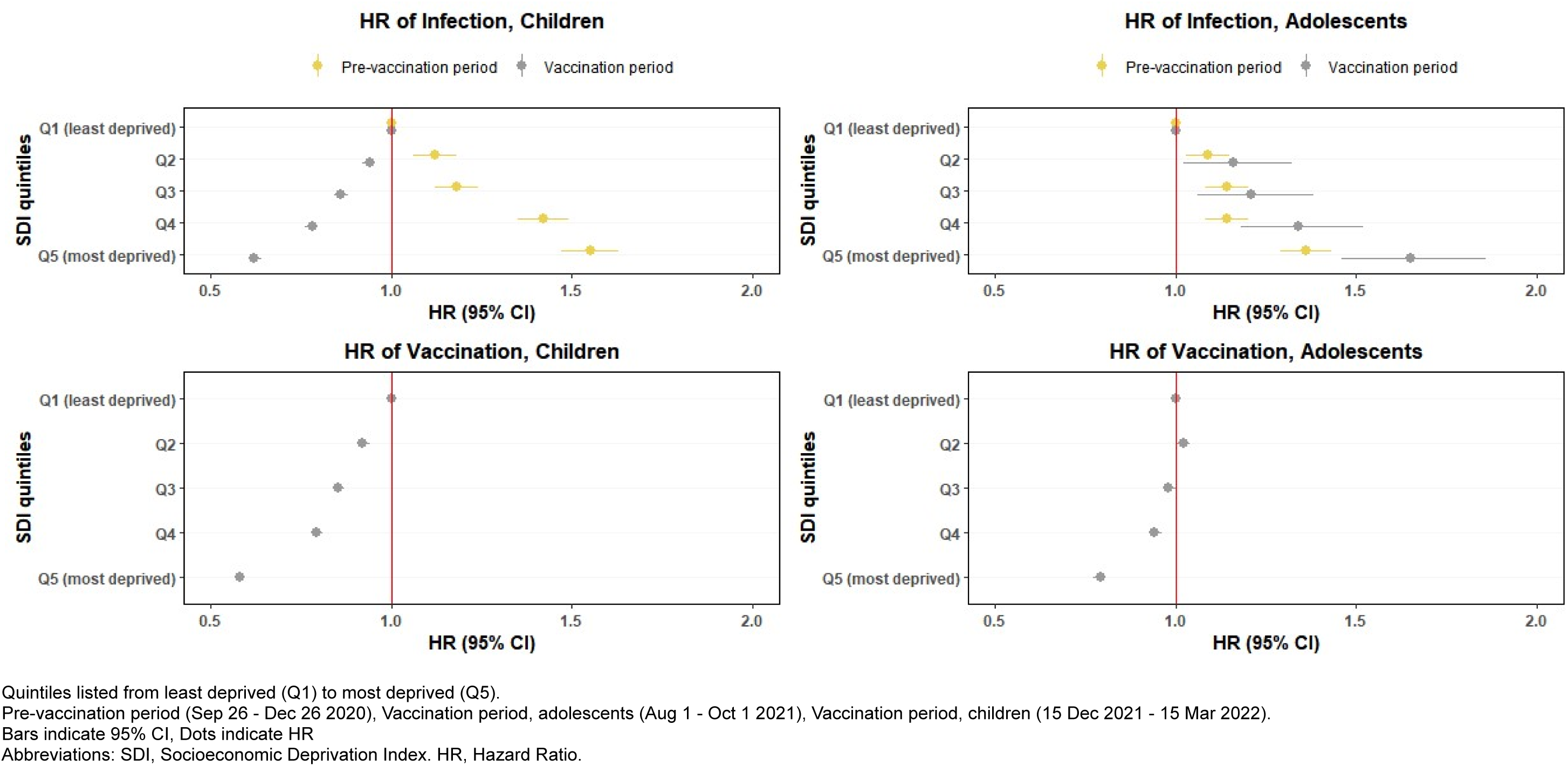
Hazard ratios of COVID-19 infection and vaccination 3 months before and 3 months after the start of the COVID-19 vaccination rollout by Socioeconomic Deprivation Index quintile.

After the vaccination rollout, COVID-19 infection risk was higher among adolescents living in more deprived areas, while in children, we observed an inverse pattern, with a lower risk of infection with more deprivation (Figure 4; eTable 4 in the Supplement). The HR of infection ranged from 1.16 (1.02 - 1.32) in Q2 areas to 1.65 (1.46 - 1.86) in Q5 areas for adolescents, and from 0.94 (0.92 - 0.95) in Q2 areas to 0.62 (0.61 - 0.64) in Q5 areas for children.

In both age groups, those living in more deprived areas had a lower risk of vaccination (Figure 4; eTable 4 in the Supplement). The HR ranged from 0.92 (0.91 - 0.94) in Q2 areas to 0.58 (0.57 - 0.59) in Q5 areas for children and from 1.02 (1.00 - 1.04) in Q2 areas to 0.79 (0.77 - 0.80) in Q5 areas for adolescents.

In our secondary analyses stratifying by sex, our results were consistent with our main analyses, showing no difference by sex (eTable 4 in the Supplement). We found similar results in sensitivity analysis after excluding individuals with previous COVID-19 (eTable 4 in the Supplement).

## Discussion

In this large population-based cohort study including 290,625 children and 179,685 adolescents, we observed a gradient of increased cumulative COVID-19 infection rates with more deprivation in children and adolescents prior to the vaccination rollout. However, over the three months following the vaccination rollout, this pattern reversed among children, with a higher proportion of infection in less deprived areas. Cumulative vaccination rates also differed by deprivation, with a gradient of increased rates with less deprivation among children and adolescents.

The association between deprivation and recorded COVID-19 infections can be explained by several factors. Previous studies on infectious diseases, including COVID-19, have demonstrated a positive correlation between disease incidence and deprivation, especially among adults^39–46^. Deprived areas typically have higher unemployment rates and lower education levels, which can increase the burden of COVID-19 cases due to comorbidities or unhealthy behaviors^47^. Moreover, lower education levels could lead to higher incidence rates of infection due to factors such as concentration in low-occupation jobs with higher exposure to precarious working conditions, or differences in health literacy^48^. While these conditions primarily affect adults, a similar trend has been observed among children, particularly within the most vulnerable populations. In the United States, for instance, racial and ethnic minorities and socioeconomically disadvantaged children have borne a disproportionate share of COVID-19 infections, as evidenced by higher rates of cases^10, 11^.This phenomenon may be attributable, in part, to intrafamily transmission dynamics^49^. Indeed, the presence of overcrowded living conditions may be a contributing factor, as they are more prevalent in economically deprived regions and have previously been associated with an elevated risk of COVID-19-related mortality^50^. It is worth noting that during the pre-vaccination period, intensive surveillance policies were implemented in schools, regardless of deprivation levels, which enabled a more accurate and equitable reflection of COVID-19 infections.

During the vaccination period in children, which spanned from December 2021 to March 2022, a higher identification of cases in less socioeconomically deprived areas was found. This could likely be attributed to differences in testing practices over time. During the vaccination period, schools had changed their protocols and stopped conducting on-site tests. Free tests were made available at pharmacies and children who had been in close contact with a positive case were advised to take a test. Although testing was readily accessible during this period, it was up to the families to take the initiative to get tested^51^. The higher incidence of recorded COVID-19 cases among children in less deprived areas aligns with the higher proportion of children in those same areas who underwent COVID-19 testing during this period.

COVID-19 vaccine uptake disparities among children are associated with several factors, including household income, neighborhood, parental education, employment, vaccination status, race and ethnicity^8, 52^. In Canada, McKinnon et al. reported that parents from lower-income households (<$100,000) had an 18.4% lower prevalence of intention to vaccinate their children than those from higher-income households (≥$150,000)^8^. Similarly, in the UK, children aged 5-11 from the most deprived quintiles had lower first-dose vaccine uptake rates than their less deprived peers (32.2% vs. 51.4%), as did those aged 12-15 (38.3% vs 68.3%)^9^. Roel et al. found similar results in when looking at the adult population aged >= 40 years from Catalonia, Spain, where COVID-19 vaccine uptake among working-age adults (40-64 years) varied by socioeconomic quintile, with higher odds of non-vaccination for persons living in more deprived areas (OR of 1.01 in Q2, and 1.33 in Q5, when compared with those living in Q1 areas)^7^. While in our study there is greater inequality in non-vaccination with deprivation among children and adolescents compared to working-age groups, both studies highlight the significant impact of area deprivation on lower vaccination rates across different age groups. Factors such as the epidemiology of the infection, vaccine prioritization strategies and parental decision-making based on attitudes, beliefs and concerns about vaccines may have contributed to these disparities^8, 53^. Overall, our results emphasize the need to address vaccine access barriers across all ages and socioeconomic levels.

The main strength of this study is that the results represent the pediatric population aged between 5 and 15 years old residing in urban areas of Catalonia, providing a comprehensive understanding of COVID-19 infection and vaccine uptake in this region of Spain. In addition, the risk of misclassification bias for vaccination is low, since SIDIAP captures all vaccines administered in Catalonia and records all COVID-19 tests performed within the public healthcare system.

This study also has limitations. First, we lacked information on individual-level SES, and therefore there is a risk of ecological bias by the potential misclassification of quintiles of SES. Secondly, deprivation was estimated based on data from the 2001 census national tract, whereas our study period spanned from 2020 to 2022. Additionally, deprivation was only available in urban areas, and therefore, our results are not generalisable to rural populations. Thirdly, herd immunity, changes in testing patterns throughout the pandemic and circulating COVID-19 variants may have affected the comparability of study periods in terms of COVID-19 infections. It is worth noting that the vaccination period partially coincided with school breaks for both children and adolescents. Although COVID-19 testing was widely available through primary care and pharmacies, holidays could have potentially affected the detection of COVID-19 cases, as testing efforts in schools were not in place during this time^23^. Lastly, we were unable to control for potential confounding variables related to the parents, such as their age or nationality.

In this study, we found a pattern of increased HR of COVID-19 vaccination with deprivation among children and adolescents. Infection inequalities persisted 3 months after vaccine rollout in adolescents. However, the situation appeared to reverse for children, possibly due to disparities in testing practices.

## Data Availability

In accordance with current European and national law, the data used in this study is only available for the researchers participating in this study. Thus, we are not allowed to distribute or make publicly available the data to other parties. However, researchers from public institutions can request data from SIDIAP if they comply with certain requirements. Further information is available online or by contacting SIDIAP.

https://sidiap.org/index.php/en/solicituds-en

## Acknowledgments

**Conflict of Interest Disclosures**

The authors declare that there is no conflict of interest.

**Funding**

This research received no specific grant from any funding agency in the public, commercial or not-for-profit sectors.

**Data sharing statement**

In accordance with current European and national law, the data used in this study is only available for the researchers participating in this study. Thus, we are not allowed to distribute or make publicly available the data to other parties. However, researchers from public institutions can request data from SIDIAP if they comply with certain requirements. Further information is available online (https://www.sidiap.org/index.php/menu-solicitudesen/application-proccedure) or by contacting SIDIAP.

**Author Contributions**

IL-S, TD-S, BR and ER conceived and designed the study. IL-S drafted the initial version of the manuscript. All authors interpreted the results, critically reviewed the manuscript and approved the final version for submission.

**Additional Contributors**

The authors would like to acknowledge the efforts of all public healthcare workers involved in the management of the COVID-19 pandemic during these challenging times in Catalonia. We also want to thank the Institut Català de la Salut (ICS) and the Programa d’analítica de dades per a la recerca i la innovació en salut (PADRIS) for providing access to the different data sources accessible through SIDIAP. We would also like to thank the staff at the IDIAP Jordi Gol who collaborated on the development of this study.

