## Supplementary material for "Socioeconomic inequalities in COVID-19 infection and vaccine uptake among children and adolescents in Catalonia, Spain": Figures and Tables

### Supplemental Online Content

**eTable 1.** Baseline characteristics of the children of Catalonia, Spain, included in the study of socioeconomic inequality in COVID-19 infection and vaccination.

**eTable 2.** Baseline characteristics of the adolescents of Catalonia, Spain included in the study of socioeconomic inequality in COVID-19 infection and vaccination.

**eTable 3.** Cumulative incidence of COVID-19 infection and vaccine coverage by cohort, overall and by sex and Socioeconomic Deprivation Index quintile.

**eTable 4.** Crude Hazard Ratios of COVID-19 infection and vaccination 3 months before and after vaccination rollout by Socioeconomic Deprivation Index quintile by SDI quintile relative to Q1, stratified by sex and previous COVID-19 infection.

**eFigure 1.** Log-log survival curves to COVID-19 infection or vaccination by Socioeconomic Deprivation Index quintile, for each study period.

**eFigure 2.** Directed Acyclic Graph modelling the relationship between socioeconomic deprivation and COVID-19 infection among children and adolescents.

**eFigure 3.** Directed Acyclic Graph modelling the relationship between socioeconomic deprivation and COVID-19 vaccine uptake among children and adolescents.

**eFigure 4.** Proportion of individuals who had performed one or more tests by age group and Socioeconomic Deprivation Index quintile, before and after the start of the vaccination rollout.

**eTable 1. Baseline characteristics of the children of Catalonia, Spain, included in the study of socioeconomic inequality in COVID-19 infection and vaccination.**

| Characteristics | Pre-vaccination period, Infection | Vaccination period, Infection | Vaccination period, Vaccine uptake |
|---|---|---|---|
| **N** | 290625 | 276753 | 274472 |
| **Age (median [IQR])** | 8.0 [6.0, 10.0] | 8.0 [6.0, 10.0] | 8.0 [6.0, 10.0] |
| **Sex No.(%)** | | | |
| Female | 141020 (48.5) | 134520 (48.6) | 133468 (48.6) |
| Male | 149605 (51.5) | 142233 (51.4) | 141004 (51.4) |
| **Nationality No.(%)** | | | |
| Spain | 250644 (86.2) | 234209 (84.6) | 232036 (84.5) |
| Africa | 12915 (4.4) | 13797 (5.0) | 13773 (5.0) |
| Asia & Oceania | 9492 (3.3) | 9441 (3.4) | 9430 (3.4) |
| Central & South America | 8802 (3.0) | 10577 (3.8) | 10544 (3.8) |
| Eastern Europe | 5487 (1.9) | 5251 (1.9) | 5225 (1.9) |
| Europe & North America | 3285 (1.1) | 3478 (1.3) | 3464 (1.3) |
| **SDI quintile No.(%)** | | | |
| Q1 (least deprived) | 53140 (18.3) | 50188 (18.1) | 49699 (18.1) |
| Q2 | 55703 (19.2) | 52301 (18.9) | 51795 (18.9) |
| Q3 | 56850 (19.6) | 53707 (19.4) | 53234 (19.4) |
| Q4 | 60099 (20.7) | 57426 (20.7) | 56973 (20.8) |
| Q5 (most deprived) | 64833 (22.3) | 63131 (22.8) | 62771 (22.9) |
| **Previous COVID-19 Infection No.(%)** | 1814 (0.6) | 47300 (17.1) | 46640 (17.0) |

*Values are no. (%) except as indicated. Pre-vaccination period (Sep 26 - Dec 26 2020), Vaccination period, children (15 Dec 2021 - 15 Mar 2022). Quintiles listed from least deprived (Q1) to most deprived (Q5). IQR, interquartile range; SDI, Socioeconomic Deprivation Index; COVID-19; Coronavirus Disease 2019.

**eTable 2. Baseline characteristics of the adolescents of Catalonia, Spain included in the study of socioeconomic inequality in COVID-19 infection and vaccination.**

| Characteristics | Pre-vaccination period, Infection | Vaccination period, Infection | Vaccination period, Vaccine uptake |
|---|---|---|---|
| **N** | 179685 | 175249 | 174807 |
| **Age (median [IQR])** | 13.0 [12.0, 14.0] | 13.0 [12.0, 14.0] | 13.0 [12.0, 14.0] |
| **Sex No.(%)** | | | |
| Female | 87243 (48.6) | 84727 (48.3) | 84516 (48.3) |
| Male | 92442 (51.4) | 90522 (51.7) | 90291 (51.7) |
| **Nationality No.(%)** | | | |
| Spain | 159824 (88.9) | 153932 (87.8) | 153552 (87.8) |
| Africa | 5247 (2.9) | 5641 (3.2) | 5623 (3.2) |
| Asia & Oceania | 5065 (2.8) | 5329 (3.0) | 5316 (3.0) |
| Central & South America | 4908 (2.7) | 5508 (3.1) | 5490 (3.1) |
| Eastern Europe | 3030 (1.7) | 3182 (1.8) | 3174 (1.8) |
| Europe & North America | 1611 (0.9) | 1657 (0.9) | 1652 (0.9) |
| **SDI quintile No.(%)** | | | |
| Q1 (least deprived) | 34524 (19.2) | 32518 (18.6) | 32440 (18.6) |
| Q2 | 35637 (19.8) | 34477 (19.7) | 34397 (19.7) |
| Q3 | 35686 (19.9) | 34788 (19.9) | 34700 (19.9) |
| Q4 | 35994 (20.0) | 35595 (20.3) | 35495 (20.3) |
| Q5 (most deprived) | 37844 (21.1) | 37871 (21.6) | 37775 (21.6) |
| **Previous COVID-19 Infection No.(%)** | 1039 (0.6) | 30323 (17.3) | 29958 (17.1) |

*Values are no. (%) except as indicated. Pre-vaccination period (Sep 26 - Dec 26 2020), Vaccination period, adolescents (Aug 1 - Oct 1 2021). Quintiles listed from least deprived (Q1) to most deprived (Q5). IQR, interquartile range; SDI, Socioeconomic Deprivation Index; COVID-19; Coronavirus Disease 2019.

**eTable 3. Cumulative incidence of COVID-19 infection and vaccine coverage by cohort, overall and by sex and Socioeconomic Deprivation Index quintile.**

| | Overall | Q1 | Q2 | Q3 | Q4 | Q5 |
|---|---|---|---|---|---|---|
| **Pre-vaccination rollout, Children** | | | | | | |
| Infection No.(%) | | | | | | |
| *Overall* | 16773 (5.8) | 2441 (4.6) | 2862 (5.1) | 3058 (5.4) | 3867 (6.4) | 4545 (7.0) |
| *Female* | 8122 (5.8) | 1174 (4.6) | 1366 (5.1) | 1488 (5.4) | 1912 (6.6) | 2182 (6.9) |
| *Male* | 8651 (5.8) | 1267 (4.6) | 1496 (5.2) | 1570 (5.4) | 1955 (6.3) | 2363 (7.1) |
| **Pre-vaccination rollout, Adolescents** | | | | | | |
| Infection No.(%) | | | | | | |
| *Overall* | 15430 (8.6) | 2597 (7.5) | 2908 (8.2) | 3039 (8.5) | 3078 (8.6) | 3808 (10.1) |
| *Female* | 7542 (8.6) | 1261 (7.5) | 1406 (8.1) | 1464 (8.4) | 1529 (8.8) | 1882 (10.3) |
| *Male* | 7888 (8.5) | 1336 (7.6) | 1502 (8.2) | 1575 (8.6) | 1549 (8.4) | 1926 (9.8) |
| **Vaccination rollout, Children** | | | | | | |
| Infection No.(%) | | | | | | |
| *Overall* | 92958 (33.6) | 19729 (39.3) | 19501 (37.3) | 18656 (34.7) | 18334 (31.9) | 16738 (26.5) |
| *Female* | 44677 (33.2) | 9436 (38.7) | 9414 (37.2) | 9079 (34.7) | 8697 (31.2) | 8051 (26.1) |
| *Male* | 48281 (33.9) | 10293 (39.9) | 10087 (37.3) | 9577 (34.8) | 9637 (32.6) | 8687 (26.9) |
| Vaccination No.(%) | | | | | | |
| *Overall* | 123018 (44.8) | 24970 (50.2) | 25117 (48.5) | 24743 (46.5) | 25465 (44.7) | 22723 (36.2) |
| *Female* | 59975 (44.9) | 12141 (50.2) | 12173 (48.6) | 11998 (46.2) | 12484 (45.1) | 11179 (36.5) |
| *Male* | 63043 (44.7) | 12829 (50.2) | 12944 (48.4) | 12745 (46.7) | 12981 (44.3) | 11544 (35.9) |
| **Vaccination rollout, Adolescents** | | | | | | |
| Infection No.(%) | | | | | | |
| *Overall* | 2815 (1.6) | 408 (1.3) | 502 (1.5) | 527 (1.5) | 598 (1.7) | 780 (2.1) |
| *Female* | 1466 (1.7) | 208 (1.3) | 266 (1.6) | 284 (1.7) | 293 (1.7) | 415 (2.3) |
| *Male* | 1349 (1.5) | 200 (1.2) | 236 (1.3) | 243 (1.4) | 305 (1.7) | 365 (1.9) |
| Vaccination No.(%) | | | | | | |
| *Overall* | 126995 (72.6) | 24216 (74.6) | 25853 (75.2) | 25716 (74.1) | 25924 (73.0) | 25286 (66.9) |
| *Female* | 61712 (73.0) | 11662 (74.3) | 12540 (75.5) | 12533 (74.2) | 12724 (74.1) | 12253 (67.5) |

| | | | | | | |
|---|---|---|---|---|---|---|
| *Male* | 65283 (72.3) | 12554 (75.0) | 13313 (74.9) | 13183 (74.0) | 13200 (72.0) | 13033 (66.4) |

Pre-vaccination period (Sep 26 - Dec 26 2020), Vaccination period, adolescents (Aug 1 - Oct 1 2021), Vaccination period, children (15 Dec 2021 - 15 Mar 2022). Quintiles listed from least deprived (Q1) to most deprived (Q5). IQR, interquartile range; SDI, Socioeconomic Deprivation Index.

**eTable 4. Crude Hazard Ratios of COVID-19 infection and vaccination 3 months before and after vaccination rollout by Socioeconomic Deprivation Index quintile by SDI quintile relative to Q1, stratified by sex and previous COVID-19 infection.**

**Infection, Pre-vaccination period, Children**

| | HR (95% CI) | HR Female (95% CI) | HR Male (95% CI) | HR Without Previous COVID-19 (95% CI) |
|---|---|---|---|---|
| *Q1 (ref.)* | 1 [Reference] | 1 [Reference] | 1 [Reference] | 1 [Reference] |
| *Q2* | 1.12 (1.06 - 1.18) | 1.12 (1.04 - 1.21) | 1.12 (1.04 - 1.21) | 1.12 (1.06 - 1.18) |
| *Q3* | 1.18 (1.12 - 1.24) | 1.18 (1.10 - 1.28) | 1.17 (1.09 - 1.26) | 1.17 (1.11 - 1.24) |
| *Q4* | 1.42 (1.35 - 1.49) | 1.46 (1.35 - 1.57) | 1.38 (1.28 - 1.48) | 1.41 (1.34 - 1.49) |
| *Q5* | 1.55 (1.47 - 1.63) | 1.54 (1.43 - 1.65) | 1.56 (1.46 - 1.67) | 1.55 (1.47 - 1.63) |

**Infection, Pre-vaccination period, Adolescents**

| | HR (95% CI) | HR Female (95% CI) | HR Male (95% CI) | HR Without Previous COVID-19 (95% CI) |
|---|---|---|---|---|
| *Q1 (ref.)* | 1 [Reference] | 1 [Reference] | 1 [Reference] | 1 [Reference] |
| *Q2* | 1.09 (1.03 - 1.15) | 1.08 (1.01 - 1.17) | 1.09 (1.01 - 1.17) | 1.09 (1.03 - 1.15) |
| *Q3* | 1.14 (1.08 - 1.20) | 1.13 (1.05 - 1.22) | 1.15 (1.07 - 1.23) | 1.13 (1.08 - 1.20) |
| *Q4* | 1.14 (1.08 - 1.20) | 1.18 (1.09 - 1.27) | 1.11 (1.03 - 1.20) | 1.14 (1.08 - 1.20) |
| *Q5* | 1.36 (1.29 - 1.43) | 1.40 (1.31 - 1.51) | 1.32 (1.23 - 1.41) | 1.36 (1.29 - 1.43) |

**Infection, Vaccination period, Children**

| | HR (95% CI) | HR Female (95% CI) | HR Male (95% CI) | HR Without Previous COVID-19 (95% CI) |
|---|---|---|---|---|
| *Q1 (ref.)* | 1 [Reference] | 1 [Reference] | 1 [Reference] | 1 [Reference] |
| *Q2* | 0.94 (0.92 - 0.95) | 0.96 (0.93 - 0.98) | 0.92 (0.89 - 0.94) | 0.94 (0.92 - 0.96) |
| *Q3* | 0.86 (0.84 - 0.88) | 0.88 (0.85 - 0.90) | 0.84 (0.82 - 0.87) | 0.86 (0.84 - 0.87) |
| *Q4* | 0.78 (0.76 - 0.79) | 0.77 (0.75 - 0.79) | 0.78 (0.76 - 0.80) | 0.78 (0.76 - 0.79) |
| *Q5* | 0.62 (0.61 - 0.64) | 0.63 (0.61 - 0.65) | 0.62 (0.60 - 0.64) | 0.62 (0.61 - 0.64) |

**Infection, Vaccination period, Adolescents**

| | HR (95% CI) | HR Female (95% CI) | HR Male (95% CI) | HR Without Previous COVID-19 (95% CI) |
|---|---|---|---|---|
| *Q1 (ref.)* | 1 [Reference] | 1 [Reference] | 1 [Reference] | 1 [Reference] |
| *Q2* | 1.16 (1.02 - 1.32) | 1.21 (1.01 - 1.45) | 1.11 (0.92 - 1.34) | 1.13 (0.98 - 1.30) |
| *Q3* | 1.21 (1.06 - 1.38) | 1.27 (1.06 - 1.52) | 1.14 (0.95 - 1.38) | 1.19 (1.03 - 1.36) |
| *Q4* | 1.34 (1.18 - 1.52) | 1.29 (1.08 - 1.54) | 1.40 (1.17 - 1.67) | 1.35 (1.18 - 1.55) |
| *Q5* | 1.65 (1.46 - 1.86) | 1.73 (1.47 - 2.05) | 1.56 (1.31 - 1.86) | 1.60 (1.41 - 1.82) |

**Vaccination, Vaccination period, Children**

| | HR (95% CI) | HR Female (95% CI) | HR Male (95% CI) | HR Without Previous COVID-19 (95% CI) |
|---|---|---|---|---|
| *Q1 (ref.)* | 1 [Reference] | 1 [Reference] | 1 [Reference] | 1 [Reference] |

| | | | | |
|---|---|---|---|---|
| *Q2* | 0.92 (0.91 - 0.94) | 0.93 (0.90 - 0.95) | 0.92 (0.90 - 0.94) | 0.92 (0.91 - 0.94) |
| *Q3* | 0.85 (0.84 - 0.87) | 0.85 (0.83 - 0.87) | 0.86 (0.83 - 0.88) | 0.85 (0.84 - 0.87) |
| *Q4* | 0.79 (0.78-0.81) | 0.80 (0.78 - 0.82) | 0.78 (0.76 - 0.80) | 0.79 (0.78-0.81) |
| *Q5* | 0.58 (0.57 - 0.59) | 0.59 (0.57 - 0.60) | 0.58 (0.56 - 0.59) | 0.57 (0.56 - 0.58) |

**Vaccination, Vaccination period, Adolescents**

| | HR (95% CI) | HR Female (95% CI) | HR Male (95% CI) | HR Without Previous COVID-19 (95% CI) |
|---|---|---|---|---|
| *Q1 (ref.)* | 1 [Reference] | 1 [Reference] | 1 [Reference] | 1 [Reference] |
| *Q2* | 1.02 (1.00 - 1.04) | 1.03 (1.01 - 1.06) | 1.01 (0.99 - 1.03) | 1.02 (1.00 - 1.04) |
| *Q3* | 0.98 (0.97 - 1.00) | 0.99 (0.97 - 1.02) | 0.98 (0.95 - 1.00) | 0.98 (0.96 - 1.00) |
| *Q4* | 0.94 (0.93 - 0.96) | 0.98 (0.95 - 1.00) | 0.91 (0.89 - 0.94) | 0.94 (0.92 - 0.96) |
| *Q5* | 0.79 (0.77 - 0.80) | 0.81 (0.79 - 0.83) | 0.77 (0.75 - 0.79) | 0.77 (0.76 - 0.79) |

Quintiles listed from least deprived (Q1) to most deprived (Q5). SDI, Socioeconomic Deprivation Index; COVID-19; Coronavirus Disease 2019. Pre-vaccination period (Sep 26 - Dec 26 2020), Vaccination period, adolescents (Aug 1 - Oct 1 2021), Vaccination period, children (15 Dec 2021 - 15 Mar 2022).

**eFigure 1. Log-log survival curves to COVID-19 infection or vaccination by Socioeconomic Deprivation Index quintile, for each study period.**

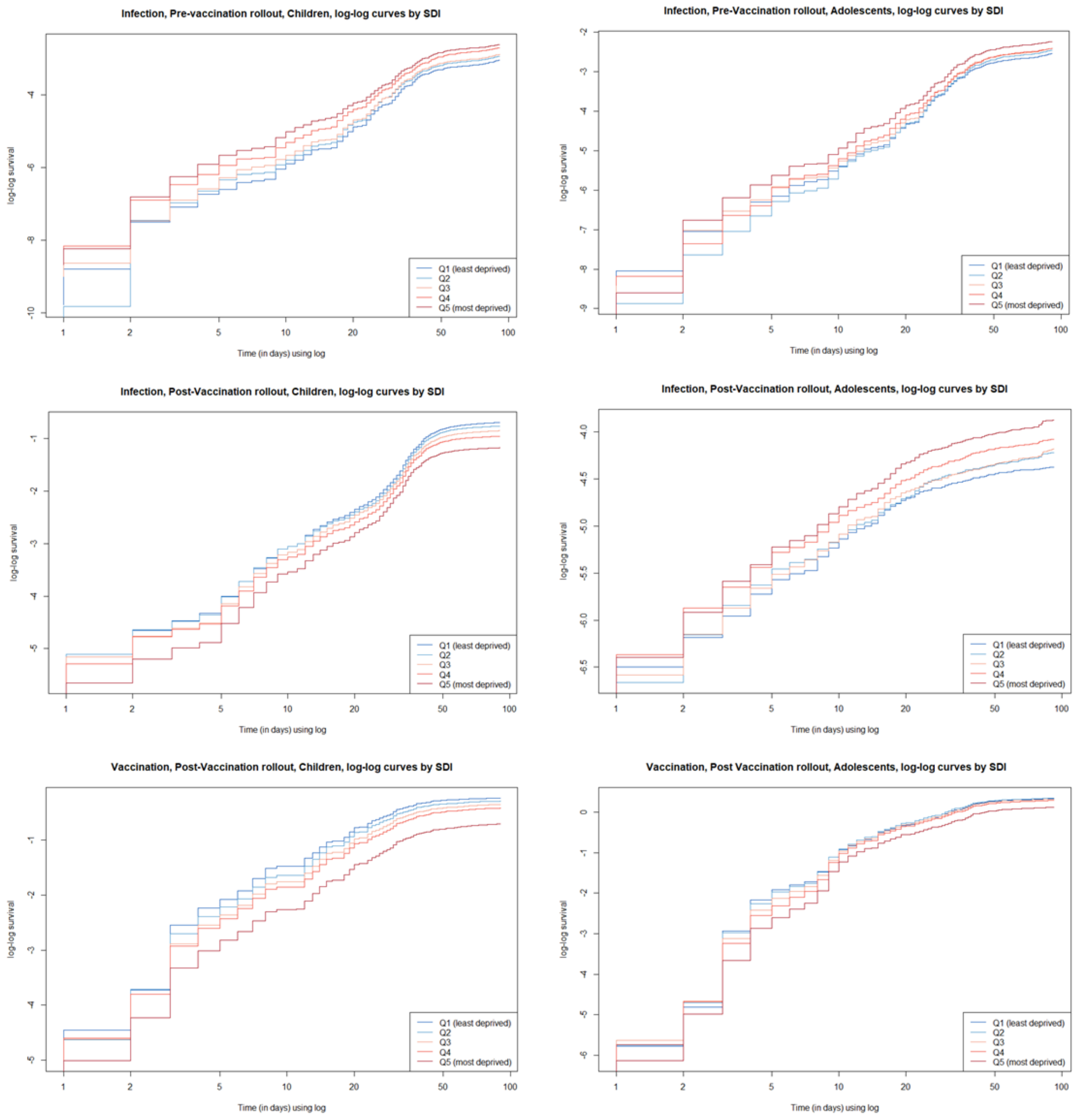


Pre-vaccination period (Sep 26 - Dec 26 2020), Vaccination period, adolescents (Aug 1 - Oct 1 2021), Vaccination period, children (15 Dec 2021 - 15 Mar 2022). Quintiles listed from least deprived (Q1) to most deprived (Q5). IQR, interquartile range; SDI, Socioeconomic Deprivation Index

**eFigure 2. Directed Acyclic Graph modelling the relationship between socioeconomic deprivation and COVID-19 infection among children and adolescents.**

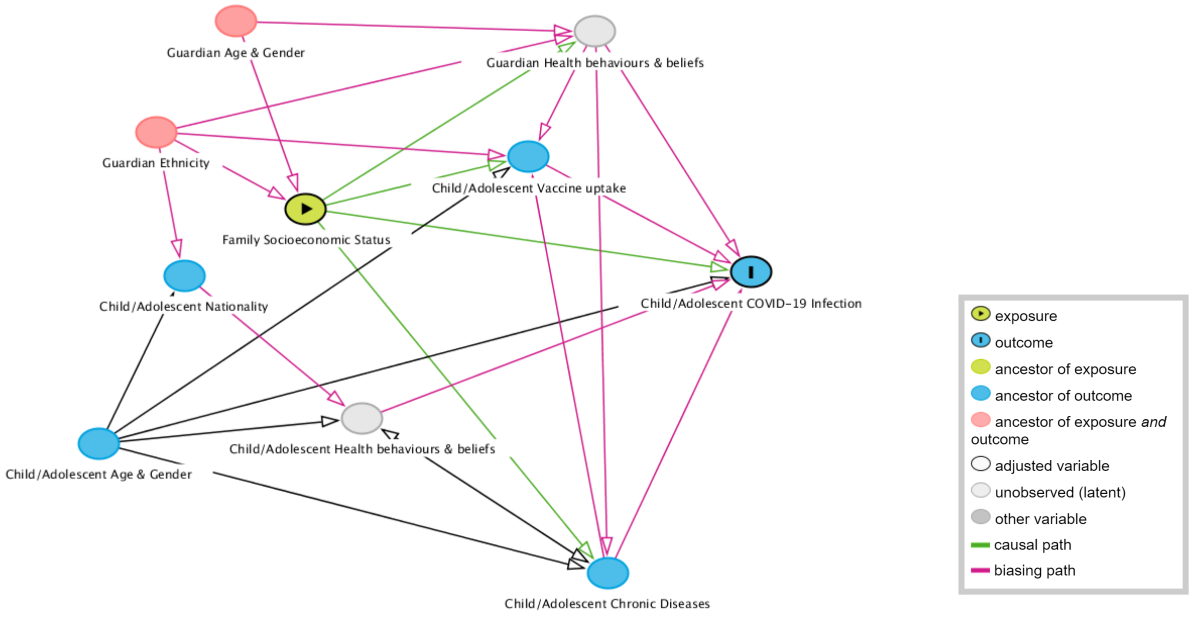


COVID-19; Coronavirus Disease 2019

**eFigure 3. Directed Acyclic Graph modelling the relationship between socioeconomic deprivation and COVID-19 vaccine uptake among children and adolescents.**

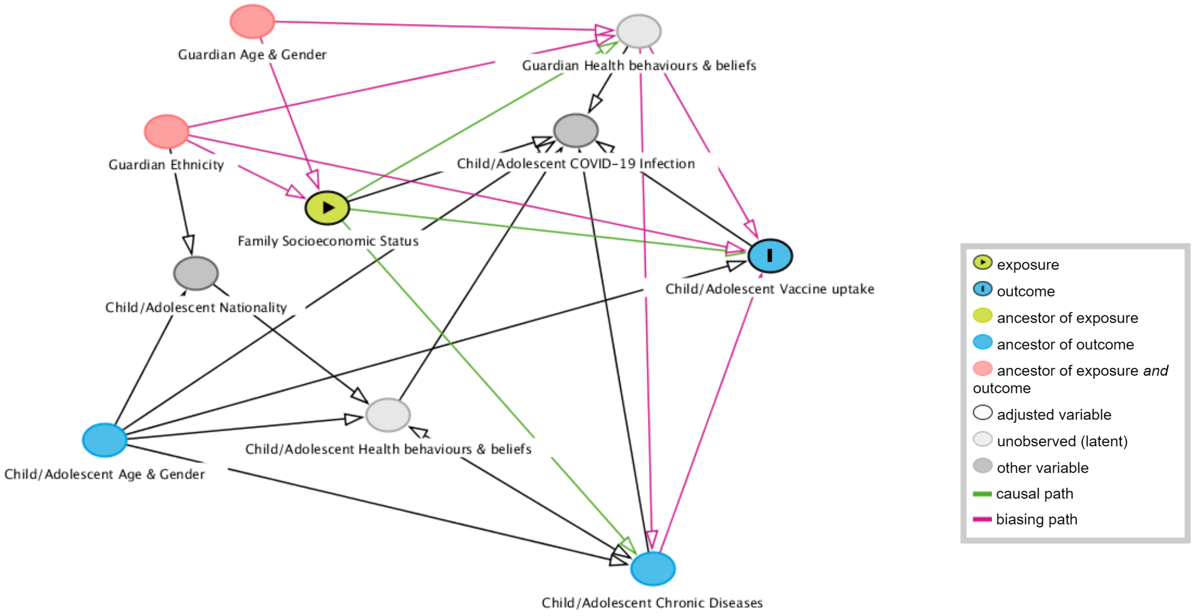


COVID-19; Coronavirus Disease 2019

**eFigure 4. Proportion of individuals who had performed one or more tests by age group and Socioeconomic Deprivation Index quintile, before and after the start of the vaccination rollout.**

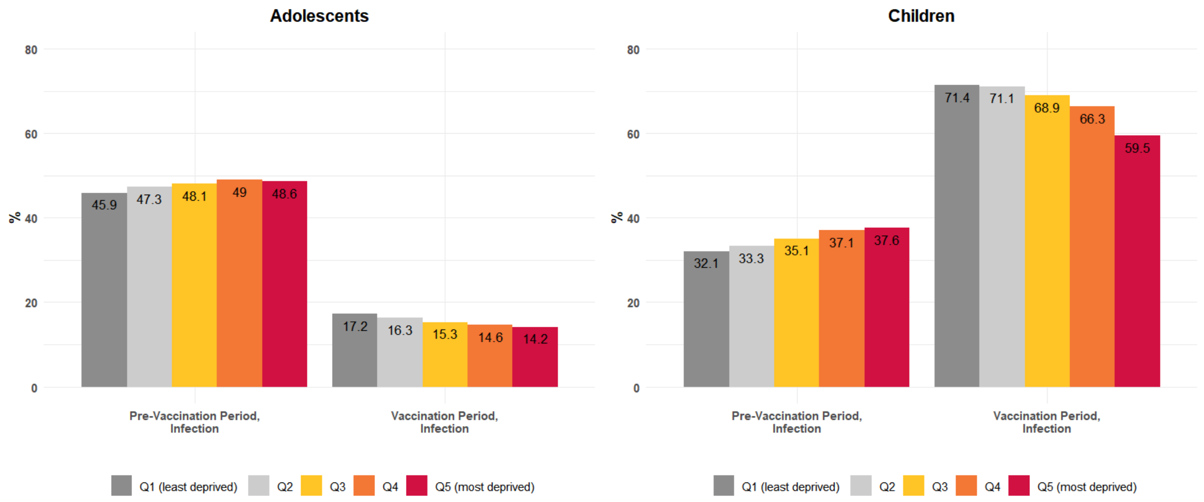


Quintiles listed from least deprived (Q1) to most deprived (Q5). SDI, Socioeconomic Deprivation Index. Pre-vaccination period (Sep 26 - Dec 26 2020), Vaccination period, adolescents (Aug 1 - Oct 1 2021), Vaccination period, children (15 Dec 2021 - 15 Mar 2022).
